# Computer Vision for Real-Time Pixel-Level Anatomical Segmentation in Neurosurgery: First-in-Human Clinical Evaluation and Iterative Development (IDEAL Stage 1)

**DOI:** 10.64898/2026.06.11.26355205

**Authors:** Danyal Z Khan, Zhehua Mao, Anjana Wijekoon, Adrito Das, Simon C Williams, Ann Blandford, Abhiney Jain, Lauren Harris, Anouk Borg, Neil Dorward, Matthew Clarkson, Sophia Bano, Peter McCulloch, Danail Stoyanov, Hani J Marcus

## Abstract

**Introduction:** Precise anatomical navigation is fundamental to safe endoscopic pituitary surgery, a high-stakes procedure characterised by a challenging learning curve. While traditional navigation systems often rely on workflow-disrupting probes or static preoperative imaging, advancements in computer vision AI (CVAI) now enable dynamic, real-time pixel-level anatomical segmentation directly from live surgical video. Our group has previously conducted a series of preclinical human-computer interaction studies to refine the system’s design, alongside digital and high-fidelity physical simulations demonstrating the benefit of AI assistance in improving overall performance, training, and safety^4–8^. Building on this foundation, the current study represents a first-in-human application of real-time pixel-level CVAI anatomical segmentation in the neurosurgical operating room, serving to assess feasibility and to iteratively improve the system.

**Method:** Guided by DECIDE-AI and IDEAL frameworks, this single-centre evaluation comprises an initial proof-of-concept phase (n=6) for endoscopic transsphenoidal pituitary surgeries. The AI model utilised a DINOv3-derived vision transformer architecture, deployed via a high-performance edge computing unit to achieve low-latency, real-time inference without reliance on cloud infrastructure. Feasibility and functionality were assessed via structured questionnaire, prospective observation, and blinded retrospective review of the recordings of the endoscopic surgical video feed and wider operating room environment. Continuous multi-stakeholder feedback through validated human factors surveys drove iterative technical refinements between cases.

**Results:** Eight patients with pituitary adenomas were enrolled. The CVAI system was successfully deployed in six cases, demonstrating acceptable real-time pixel-level sella segmentation accuracy. Deployment failed pre-operatively in two cases owing to a single recurring system reboot bug. Iterative refinement between cases were driven by our experience and surgical team feedback. This resulted in the integration of additional anatomical structure segmentations (e.g., carotid arteries), enhanced model accuracy via training dataset expansion, and hardware firmware upgrades. Multi-stakeholder surveys demonstrated satisfactory system feasibility, usability, and acceptability among the surgical team. Both prospective observation and retrospective video review confirmed the absence of adverse events, including no significant distraction to the primary surgeon, and there were no AI-related clinical complications.

**Conclusion:** This first-in-human early clinical evaluation demonstrates the feasibility and iterative development of real-time pixel-level CVAI-based anatomical navigation during high-stakes neurosurgery. Future work will include a larger single-centre case series (IDEAL Stage 2a) with more surgical teams to further iterate the system and explore its impact on safety, training and workflow. As the underpinning AI models improve and integrate with other intra-operative navigational technologies, such tools will likely be the cornerstone of intra-operative surgical decision support systems.

## Background

The mastery of anatomical knowledge and precise intra-operative navigation constitute the fundamental tenets of safe surgical practice. Inadequacy in anatomical orientation or navigational errors represent significant risks to patient safety, with the potential for serious morbidity or mortality. Consequently, a variety of surgical adjuncts have been developed to assist with orientation and navigation (e.g., image-guidance). When integrated correctly into the surgical workflow, these tools serve a dual purpose: providing real-time guidance and facilitating the consolidation of anatomical learning and reflection-in-practice.

Recent advancements in artificial intelligence (AI) and computational power have catalysed the emergence of computer vision AI (CVAI) analysis of live surgical video as a candidate technology to assist with anatomical navigation. Unlike traditional navigational systems that often rely on workflow-disrupting specialized probes or static preoperative imaging, CVAI-based analysis of live surgical video offers a dynamic alternative that can adapt to changing surgical scenes. Pixel-wise segmentation, a subtype of CVAI, offers the potential for precise localisation of surgical structures, and therefore has more potential to assist with navigation, when compared to CVAI models which focus on simple detection or classification. While the proof of concept for real time CVAI segmentation of anatomical structures has recently been established in laparoscopic general surgery^1–3^, its application in more complex, anatomically dense environments remains an active area of investigation.

Endoscopic pituitary surgery represents a compelling candidate for the clinical translation of CVAI-based navigation. From a clinical perspective, the procedure is characterized by high stakes, a steep learning curve, and a surgical field crowded with critical neurovascular structures. From an AI engineering standpoint, the entirely endoscopic nature of the procedure provides a high-quality video feed and offers a balance between workflow homogeneity and scene heterogeneity. Leveraging this, CVAI models have been developed capable of accurate anatomical segmentation^4–10^. Preclinical data indicates these models improve anatomical orientation among surgeons and residents and enhance overall performance in high-fidelity simulations^11, 12^.

The next stage in translation of CVAI-based anatomical segmentation for navigation assistance in endoscopic pituitary surgery is early clinical evaluation. However, prior to translation from the digital world to the operating room, the surgeon-AI interface must be carefully designed to maximise usability, reduce distraction and cognitive overload, and improve the surgeon’s ability to interpret AI recommendations appropriately^13^. To address this, emerging evidence has explored the optimization of AI via augmented reality overlays via digital simulations and real-time physical simulations^11, 14, 15^. The resulting optimized user-interface not only retains the previously realised performance benefits with AI-assistance, but also may improve trust calibration, a key consideration in the DECIDE-AI guidelines for early clinical translation of AI systems^16, 17^.

Thus, the goal of this clinical trial is to assess the feasibility of this CV-based anatomical segmentation during live endoscopic pituitary surgery. Adhering to the DECIDE-AI and IDEAL frameworks for surgical innovation, we employ an iterative development process guided by real-world intra-operative experience and structured feedback from the multi-stakeholder surgical team^17, 18^. To our knowledge, this study is the first published clinical case series of the application of real-time pixel-level CVAI anatomical segmentation in live neurosurgery.

## Methods

### Overview

The overarching goals of this early clinical evaluation are to iteratively improve the technology and assess its feasibility. Guided by the IDEAL Stage 1 and DECIDE-AI frameworks, a non-comparative, single-centre, proof-of-concept study was performed^17, 18^. The study was registered on a clinical trials database (NCT07568366; registered 5^th^ May 2026), ethical approval was granted via regional ethics committee (IRAS 127474) and informed consent was received from each participant accordingly.

### System Overview

The CVAI model was developed using retrospectively collected surgical video data from 111 patients who underwent endoscopic transsphenoidal pituitary surgery at the National Hospital for Neurology and Neurosurgery (NHNN)^4–6^. From these procedures, 1704 image frames were selected from surgical stages where intraoperative anatomical guidance is considered most clinically relevant, including sellotomy, durotomy, and tumour resection, based on previously published international workflow consensus recommendations^19^. Selected images were annotated using semantic segmentation of clinically relevant anatomical structures, including the sella, carotid arteries, optic nerves, and clival recess. Annotations were performed by two neurosurgical residents (DZK and LH) with experience in pituitary surgery and subsequently spot-verified by an attending neurosurgeon (HJM). A DINOv3-derived vision transformer architecture was chosen and demonstrated segmentation performance considered sufficient for prospective clinical feasibility evaluation, achieving a mean Intersection-over-Union (IoU) of 76.5% for sella segmentation on the retrospective hold-out evaluation dataset.

For intraoperative deployment, the CVAI model was integrated into the NVIDIA Holoscan sensor processing platform (NVIDIA Corporation, Santa Clara, CA, USA). The inference pipeline was optimised using NVIDIA TensorRT acceleration and deployed on a high-performance edge computing unit (NVIDIA IGX Orin Developer Kit equipped with an NVIDIA RTX 6000 Ada Generation GPU), enabling low-latency, real-time intraoperative inference without dependence on external network connectivity or cloud infrastructure, which is often underdeveloped in hospital systems [Figure 1]. This computing system was seamlessly integrated with existing operating theatre equipment – with the live endoscopic video feed from the surgical stack (Karl Storz, Tuttlingen, Germany) captured via an AJA KONA HDMI capture card, setup to use Remote Direct Memory Access to bypass the CPU and transfer video data directly into the GPU memory of the NVIDIA IGX Orin. This results in high-throughput, low-latency video display. Processed frames with AI-generated anatomical overlays were output to the existing secondary display monitor of the surgical stack in real time, with an end-to-end processing latency of 50-100ms.

**Figure 1.**
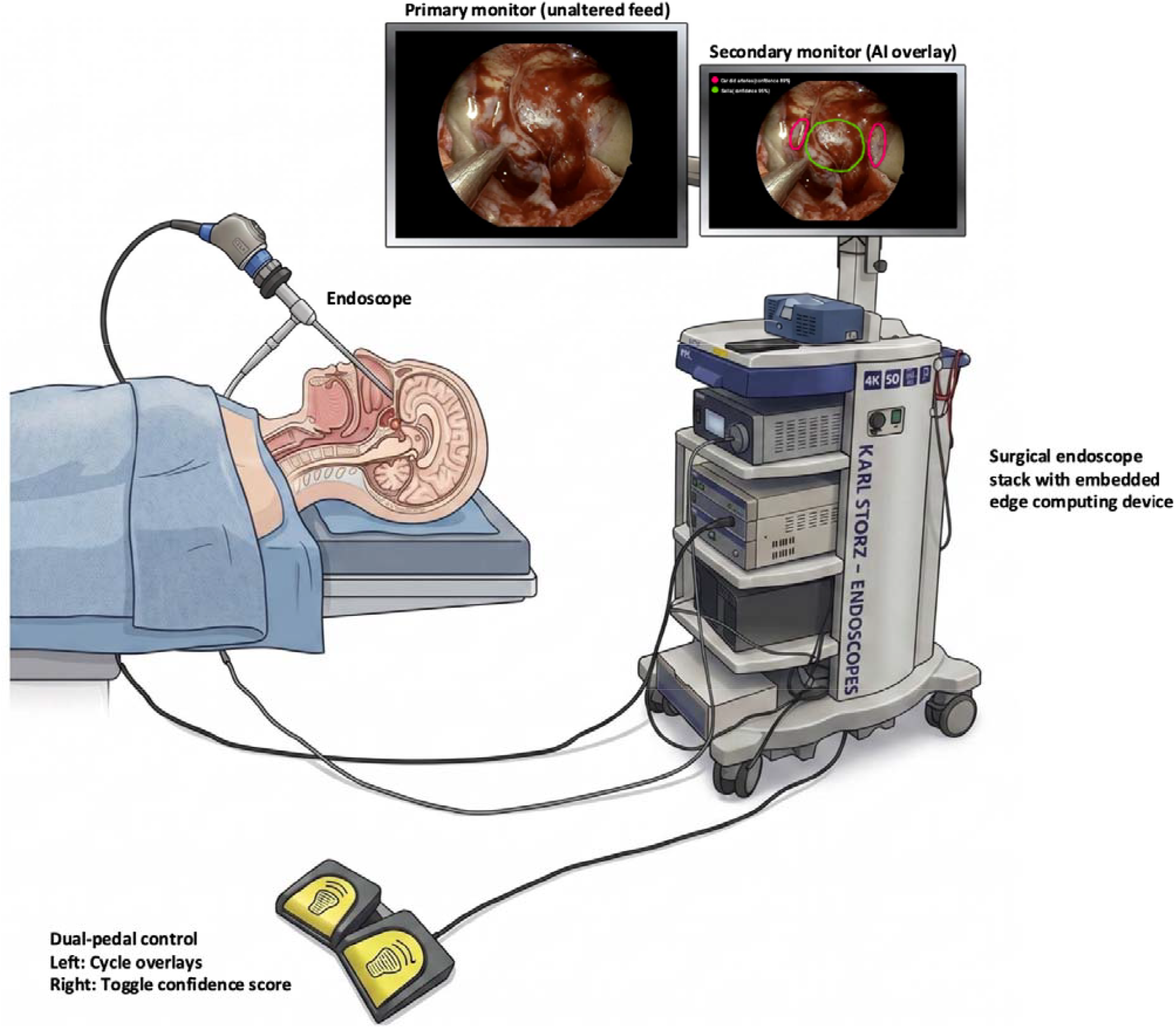
System set-up, with dataflow from the primary monitor into the NVIDIA IGX Orin computer, rather than from the video processing device, to avoid compromising the primary surgical feed. This is followed by real-time AI analysis by the deployed model in the IGX, with display of the IGX output on the in-built secondary monitor of the endoscope stack. This monitor is otherwise largely unused in our surgical practice (see IDEAL Stage 1 section). Figure made with assistance of Gemini Pro.

Importantly, the surgeon-AI interface was designed based on prior surgeon feedback during structured usability and interpretability digital simulations and real-time physical simulation studies^11, 12, 14, 15, 20^. The interface consisted of three key design features: (1) predicted pixel-level anatomical structures were rendered as outlines rather than filled masks, balancing informational salience with cognitive load; (2) activation of the CVAI overlay was placed under full surgeon control via a foot pedal, ensuring no unsolicited display of CVAI output during the procedure; and (3) a quantitative confidence metric, reflecting the model’s predictive certainty for sella segmentation, was displayed alongside the overlay to support informed intraoperative interpretation.

### Proof-of-concept study procedure

The aim of this study is demonstrating proof-of-concept, assessing feasibility, identifying safety concerns and gathering surgical team feedback to drive iterative development of the technology. In line with IDEAL Stage 1 recommendations, the sample was a small, pre-specified development cohort rather than a powered sample. Therefore, six adult patients undergoing elective endoscopic transsphenoidal surgery for pituitary adenomas were planned for enrolment. Exclusion criteria included: recent revision surgery (within 6 months), age < 18 years old, or surgeries requiring non-standard approaches (i.e. cavernous sinus exploration or extended endonasal surgery). All cases were performed at NHNN by a single attending surgeon (HJM) and their team. This team was orientated to the system using a high-fidelity simulator, with a standardised briefing on intended use, output interpretation, and the limitations of the system.^11^. In the operating room, surgery was performed as per routine using the primary unchanged surgical display, with the AI-assistance system available on the secondary monitor. This AI-enabled display remained off by default until activated by the surgical team using the foot pedal during anatomical orientation steps^19, 21^. Immediately before each case, a standardised functionality check was undertaken to confirm correct video input, system start-up, overlay rendering, user control operation, and acceptable real-time responsiveness. Cases in which the system did not pass this check proceeded without investigational deployment.

### Data collection, analysis and system iteration

Primary outcomes, measured per case, included:

1. Demonstrating proof-of-concept (system functioned as intended, prediction latency, surgeon-AI alignment on anatomical structure location, system used by user as intended): Measured via intra-operative observation and structured note taking (by a computer science postdoctoral research fellow) and video recording of the surgical video feed, AI overlays and operating theatre (via the Medtronic TouchSurgery and LiveStream ecosystem).
2. Assessing feasibility of deployment: measured via Feasibility of Intervention Measure questionnaire (FIM) and an open-ended question on how to improve it - judged by surgical team members who interacted with the system^22^.
3. Observation of safety: monitoring for primary surgeon distraction and surgical team workflow disruption (measured via event counting on review of operating room video recording) and deviation of clinical performance from the mean of a matched retrospective cohort (measured via blinded OSAT and operative time assessment of recorded surgical video feed and clinical outcome data)^23–25^.

Secondary outcomes were also assessed per case, by those who interacted with the system (surgeons & nurses), and included: 1) educational yield (measured via a structured questionnaire based on the Kirkpatrick model); 2) wider user feedback (via Acceptability of Intervention Measure, AIM; System Usability Scale, SUS; Short Form Trust in Automation Scale, STIAS, structured questionnaires)^22, 26–28^.

Baseline characteristics of the patient (age, sex, tumour type and size) and of the surgical team members that interacted with the system (age, sex, level of training, AI literacy via the Meta AI Literacy scale short form; MAILS) were collected^29^. Clinical outcomes were collected via our existing internal prospectively maintained database, which aligns with a previous multicentre collaborative study on patient outcomes. It includes length of stay, dysnatraemia, pituitary hormone dysfunction, cerebrospinal fluid leak, visual loss, resection completeness, and biochemical remission (functioning tumours only)^30^.

Data analysis included summary statistics of quantitative data (surveys and video analysis) and thematic analysis of qualitative data (observational notes and qualitative survey data) into actionable system iteration items, organised into AI and software, hardware, user interface, and intra-operative implementation domains. All changes made were logged and AI models were version controlled. Summary statistics were generated for baseline characteristics, wider user feedback and clinical outcomes.

## Results

### Overview

Eight consecutive patients meeting the inclusion criteria were recruited to the study, across five operating lists. Successful proof-of-concept of the CVAI system was demonstrated in six cases [Figure 2], with failure pre-operatively in two cases (case 2 and 3, on the same day, due to the same computer reboot bug). Four surgical team members interacted with the system across these cases – three surgeons (1 attending, 1 senior fellow, 1 resident) and one scrub nurse (with weekly pituitary surgery exposure). Baseline characteristics of the group were as follows: age (3/4 35-44 years old, 1/4 25-34 years), sex (3 males, 1 female), and AI literacy (MAILS median 6.1, IQR 5-7.1; max possible score of 10). Meaningful use of the system was limited to the sellar phase, as intended, during anatomical orientation just prior to the sellotomy and durotomy steps, when other navigational assisting tools (image guidance and doppler) are routinely used.

**Figure 2.**
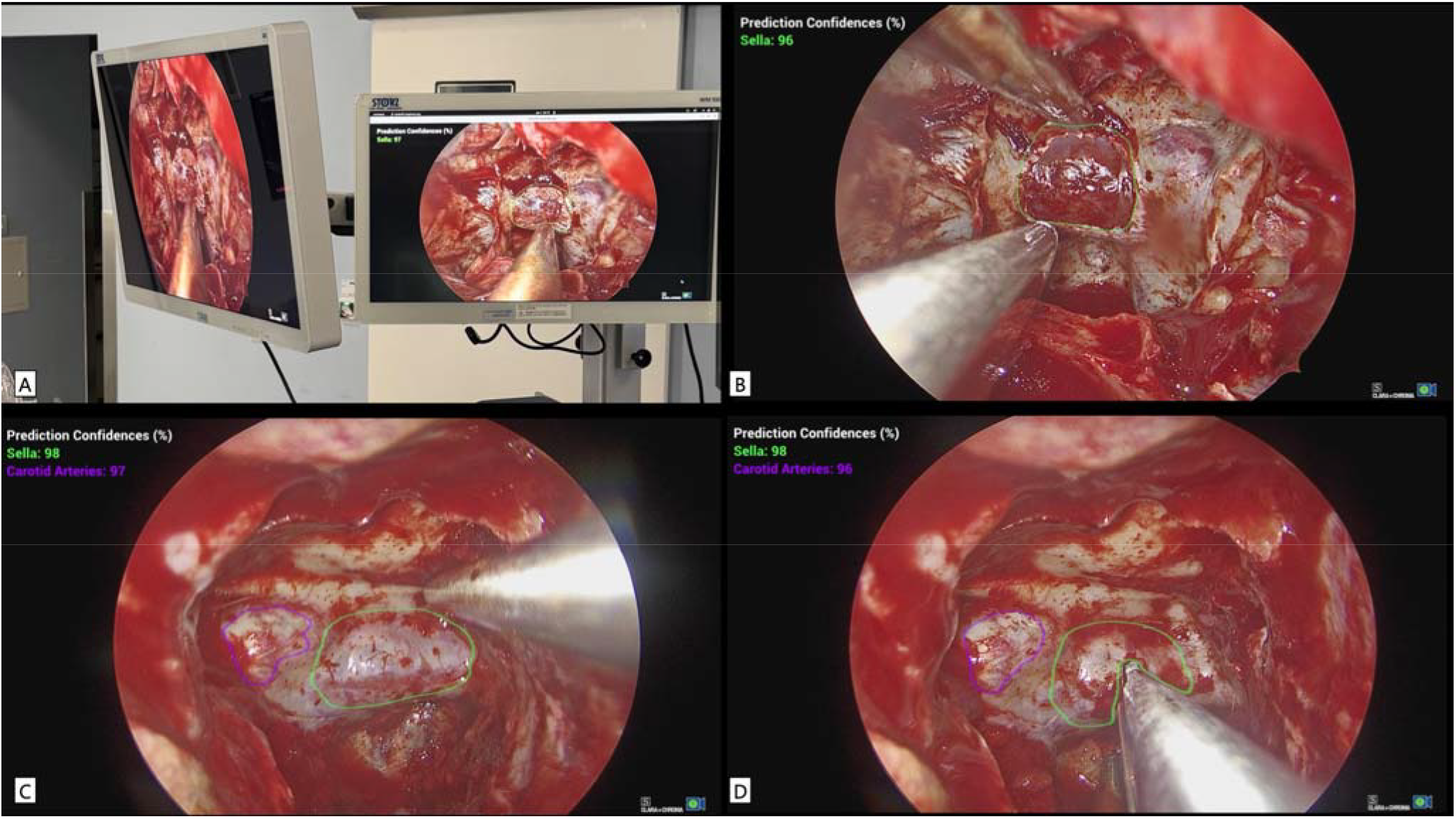
Deployment of the system in real time during endoscopic pituitary surgery. A: Intra-operative deployment. B: First case with sella-only segmentation. C: With addition of carotid artery segementation. D: Distortion of overlay by instrument.

### Feasibility

Feasibility was high across the cases series, with the median of mean participant FIM scores at 4.8 (IQR 4.3-4.8; max possible score of 5) [Table 1], with the only recommendation to improve feasibility being to deploy the model directly through existing surgical stacks rather than via a separate computer, which is currently not possible due to stack manufacturer configuration.

**Table 1.** Overview of proof of concept, feasibility and safety metrics. NFPA = non-functioning pituitary adenoma; GTR = gross total resection; STR = subtotal resection; SIADH = syndrome of inappropriate ADH. Colour coding as per operating session number. * changes between AI model versions are described in Figure 3.

| Case | Operating session | Clinical context | CVAI use | Accuracy & surgeon-AI alignment | Noticeable Latency | Median Feasibility (FIM) Score | Events of operating surgeon distraction | Events of workflow disruption | Surgical performance (mean sellar phase OSAT) | Clinical Outcomes |
| --- | --- | --- | --- | --- | --- | --- | --- | --- | --- | --- |
| 1 | 1 | 40s male<br>NFPA<br>Macroadenoma | Yes; sellar phase | AI Model V1*:<br>Sella only<br>False positive over | No | 4.25<br>(IQR 4.1-4.6) | 0 | 0 | 29/30 | No surgical complications.<br>GTR achieved. |
| 2 | 2 | 20s female<br>Acromegaly<br>Macroadenoma with cavernous sinus invasion | No; reboot malfunction | N/A | N/A | N/A | N/A | N/A | 26/30 | No surgical complications.<br>SIADH requiring treatment.<br>Cure achieved. |
| 3 | 2 | 50s female<br>NFPA<br>Macroadenoma | No; reboot malfunction | N/A | N/A | N/A | N/A | N/A | 28/30 | No surgical complications.<br>GTR achieved. |
| 4 | 3 | 20s female<br>Cyclical Cushing's disease | Yes; sellar phase | AI Model V2:<br>Sella accurate, only centre carotid prominence segmented | No | 4.5<br>(IQR 4.1-4.6) | 0 | 0 | 28/30 | No surgical complications.<br>Cure assessment requires follow-up. |
| 5 | 4 | 70s female<br>NFPA<br>Macroadenoma with cavernous sinus invasion | Yes; sellar phase | AI Model V2.1:<br>Sella & carotid satisfactory | No | 4.9<br>(IQR 4.8 4.9) | 0 | 0 | 26/30 | No surgical complications.<br>Optic decompression achieved via planned STR.<br>SIADH requiring treatment. |
| 6 | 4 | 40s male<br>Recurrent Cyclical Cushing's disease with two previous transsphenoidal surgeries | Yes; sellar phase | AI Model V2.1:<br>Sella & carotid only partially correct | Yes |  | 0 | 0 | 27/30 | No surgical complications.<br>Cure assessment requires follow-up. |
| 7 | 5 | 50s female<br>Recurrent NFPA<br>Macroadenoma with cavernous sinus invasion | Yes; sellar phase | AI Model V2.2:<br>Only sella visible, centrally accurate despite distorted anatomy | No | 4.8<br>(IQR 4.8 4.8) | 0 | 0 | 26/30 | Optic decompression achieved via planned STR.<br>No surgical complications |
| 8 | 5 | 40s male<br>NFPA<br>Giant macroadenoma | Yes; sellar phase | AI Model V2.2:<br>Sella accurate, carotid more accurate and consistent than last session | No |  | 0 | 0 | 27/30 | GTR achieved.<br>No surgical complications |

### Safety

There were no events of significant primary surgeon distraction or surgical team workflow disruption in the current implementation (Table 1). Anecdotally, there was increased attention and curiosity toward the device in the first trial (likely due to novelty) and on the fourth trial (when the carotid segmentation was added).

Surgical performance review via OSATS revealed a mean score of 27.2/30 ±1 in the sellar phase, which did not meaningfully decline from the retrospective overall score and sellar phase score of 28.6/30 ±0.8 (Supplementary Material 1))^24^. There were no intra-operative complications in this case series – including vascular or neural tissue injury. The median length of stay was 4 days. Three patients experienced anterior pituitary hormone deficiency requiring supplementation prior to discharge, and zero patients experienced posterior pituitary deficiency requiring supplementation. Two patients experienced hyponatraemia requiring treatment (case number two with no CVAI used, case number five with CVAI use). Both Cushing’s disease cases were cyclical Cushing’s and therefore assessment of remission will require longer term follow-up. Again, these were in line with retrospective cohort data (Supplementary Material 1)^24^.

### Device iteration

From surgical team feedback, intra-operative observation and surgical video review, problems were identified and addressed in a targeted fashion, as shown in Figure 3. Solutions suggested after the last operating session (operating session 5) will be attempted prior to IDEAL Stage 2a.

**Figure 3.**
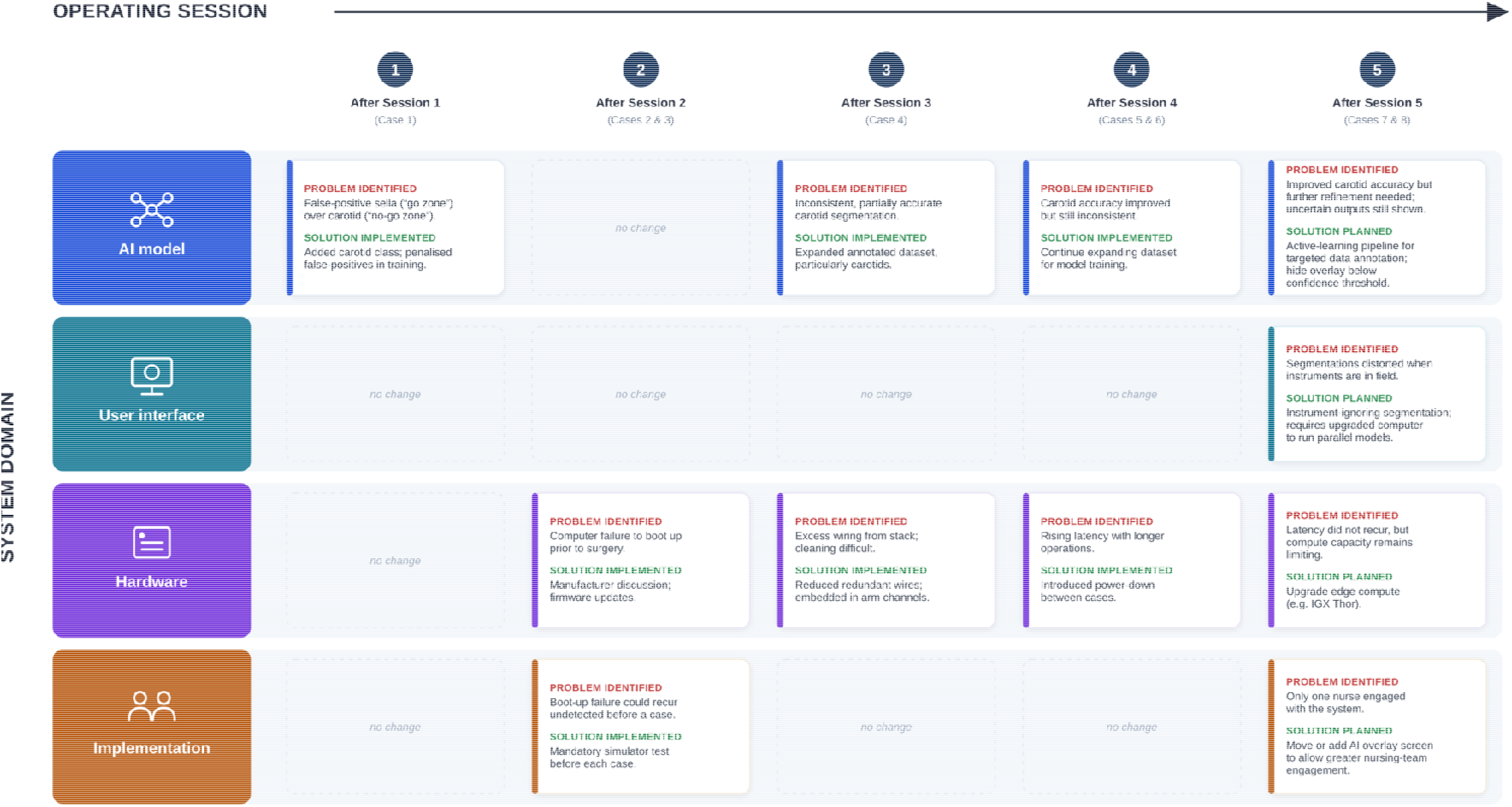
Overview of iterative changes made between operating sessions to the system. Figure aesthetics improved with assistance of Claude Opus 5.

### Wider implementation feedback and human factors

All participants saw potential as a valuable educational tool, particularly for surgeons and nurses in training. Acceptability was high across the cases series, with the median of mean participant AIM scores at 4 (IQR 3.8-4.2; max possible score of 5), with critique directed toward AI model accuracy at carotid arteries and at occlusion or distortion of AI overlay when instrument in field. Usability was also moderately high, with a SUS median score of 78 (IQR 65-79; max possible score of 100). Early trust in the system was moderate with a STIAS median of 4.3 (IQR 3.8-4.9; max possible score of 7).

## Discussion

### Principal Findings

This first-in-human IDEAL Stage 1 clinical evaluation demonstrates that deploying a real-time pixel-level CVAI anatomical segmentation system during endoscopic pituitary surgery is feasible, and early-stage implementation limitations were largely correctable. The system was successfully deployed in six of the eight planned cases, providing real-time sella and carotid pixel-level segmentation without introducing measurable distraction to the primary surgeon or disrupting operating room workflow.

The translation of CVAI from digital and simulated environments to the live neurosurgical operating room presents significant logistical and technical challenges. Iterative system development across the full ecosystem (AI model, user interface, computing hardware and implementation) was driven by deployment experience and user feedback during this case series of standard and more challenging edge-cases. Most notably, the initial model generated false-positive sella (“go-zone”) segmentations over the carotid (“no-go”) regions and exhibited overlay distortion when surgical instruments occluded the operative field. Instrument occlusion and class imbalances are widely recognized as persistent challenges in real-time surgical segmentation, frequently compromising boundary-aware metrics during in vivo deployment^31, 32^. By moving beyond dataset-level AI accuracy metrics, and instead examining clinically significant and reproducible AI errors, we have employed a targeted and iterative strategy which progressively reduced these errors as the case series progressed.

### Findings in the Context of the Literature

The current literature in surgical data science predominantly features retrospective AI models reporting high segmentation accuracies on curated, idealized datasets^31, 33^. Recent evidence suggests only 12% (n=13) of surgical CVAI studies, report the real-time capability necessary for live deployment in operating rooms^31^. Only a handful of studies have reached prospective early clinical stages (IDEAL Stage 1 & 2a), but mostly for general surgical applications - for example, laparoscopic cholecystectomy (anatomical segmentation of the “critical view of safety”) and during laparoscopic colorectal surgery (nerve recognition)^1–3^.

CVAI has been deployed in the form of bounding box based detection during pituitary surgery^34^, however, this older technique does not precisely localise surgical structures at a pixel-level, and therefore has limited impact potential for navigation and training. CVAI models that perform pixel-level segmentation are more difficult to train and are more computationally intensive, but offer the potential for precise surgical structure localisation, and thus harbour more potential for clinical impact. However, pixel-level anatomical segmentation CVAI technology is yet to be translated in live neurosurgery, a specialty characterised by the potential for significant morbidity and mortality^31^. Translation in such high stakes environments requires a proportionally rigorous approach to development and evaluation^35^. In this work, we have performed preceding multifaceted preclinical evaluations, and now present a systematic iterative approach to clinical evaluation, which goes beyond crude AI model accuracy metrics and includes deliberate user-centred design and human factors analysis^11–16, 20^. This is reflected in the principles of the IDEAL and DECIDE-AI frameworks, and thus this present study demonstrates an exemplar for the systematic preclinical development and early clinical evaluation for higher stakes surgeries – such as most neurosurgery and cardiothoracic procedures^17, 18^. For such applications, it is unlikely that CVAI will operate in isolation, and future generations should integrate further with the wider operating room. For example through multimodal analysis of video, image-guidance and other adjuncts, when AI model and computing advances eventually allow this.

### Strengths and limitations

The strengths of this study are the adherence to DECIDE-AI and IDEAL guidelines, with a multifaceted evaluation of system deployment, including human factors analysis, with the transparent reporting of iterative system failures and necessary improvements. Limitations include the early stage of this clinical evaluation (single centre, single surgical team, small case series). Furthermore, given the high-risk nature of pituitary surgery and the early stage of CVAI integration, the system was initially deployed as a educational tool rather than a primary decision-support system. Future iterations will provide direct intra-operative decision support, contingent upon the system meeting benchmarks for accuracy and consistency. Continual expansion of the training dataset (particularly for structures which are not always readily visible such as the carotid arteries, and for frames where instrument occlusion occurs) and improvements to model architecture will be key development areas. Once addressed, future work will necessitate larger case series (IDEAL Stage 2a) and prospective multicentre evaluations (IDEAL Stage 2b) to further improve the technology and implementation, and explore its benefit and risk profile at scale.

## Conclusion

This first-in-human evaluation establishes that real-time, pixel-level, edge-computed CVAI anatomical segmentation can be feasibly deployed in the neurosurgical operating room without measurable distraction to the operating surgeon, disruption to team workflow, or AI-attributable complications across the deployed cases. The iterative design of this study identified correctable failure modes which were addressed between operating sessions through targeted software, hardware, interface, and implementation changes. The next steps are a larger single-centre case series with additional surgical teams (IDEAL Stage 2a) to confirm reproducibility and refine the system further, followed by multicentre evaluation (IDEAL Stage 2b. As the underpinning AI models improve and integrate with other intra-operative navigational technologies, such tools will likely be the cornerstone of future intra-operative surgical decision support systems.

## Data availability

Upon reasonable request.

## Supplementary Material

Supplementary Material 1: Comparative analysis with retrospective cohort. Given the small prospective sample size, no formal statistical hypothesis testing was performed. OSATS: Objective Structured Assessment of Technical Skills

| Outcome measure | This study<br>(8 recruited) | This study<br>(6 CVAI-deployed) | Retrospective cohort<br>(Khan et al, n=63)<sup>24</sup> |
| --- | --- | --- | --- |
| **Surgical performance** |  |  |  |
| Mean sellar phase OSATS (/30) | 27.1 ± 1 | 27.2 ± 1 | 28.6 ± 0.8 |
| **Surgical outcomes** |  |  |  |
| Intra-operative vascular injury | 0/8 (0%) | 0/6 (0%) | 0/63 (0%) |
| Visual decline | 0/8 (0%) | 0/6 (0%) | 0/63 (0%) |
| New anterior pituitary hormone deficiency requiring supplementation | 3/8 (38%) | 2/6 (33%) | 26/63 (41%) |
| New posterior pituitary deficiency requiring supplementation | 0/8 (0%) | 0/6 (0%) | 2/63 (3.2%) |
| Hyponatraemia / dysnatraemia requiring treatment | 2/8 (25%) | 1/6 (17%) | 7/63 (11%) |
| Postoperative CSF leak | 0/8 (0%) | 0/6 (0%) | 4/63 (6.4%) |
| Length of stay, median (days) | 4 | 4 | 4 |

Supplementary Material 2: DECIDE-AI Checklist

